# The incidence of delirium in an acute geriatric community hospital: an observational cohort feasibility study

**DOI:** 10.1101/2021.05.31.21257974

**Authors:** Marthe E. Ribbink, Emma Stornebrink, Remco Franssen, Annemarieke de Jonghe, Janet L. MacNeil Vroomen, Bianca M. Buurman, on behalf of the AGCH-study group.

## Abstract

**Objectives:** Delirium in hospitalised older adults is associated with negative health outcomes. Admission to an alternative care setting may lower the incidence of delirium. The Acute Geriatric Community Hospital (AGCH) was recently opened in the Netherlands and uses a multi-component non-pharmacological intervention strategy to prevent delirium. The objective of this study was to describe the incidence of delirium at the AGCH and compare this incidence to existing rates from literature. If a possible effect on delirium is seen in this comparison this would support conducting a larger prospectively controlled study on delirium in this new care setting.

**Design:** Prospective cohort feasibility study; exploratory meta-analysis of proportions.

**Setting and Participants:** The AGCH is an acute geriatric unit in a skilled nursing facility for patients aged >65 years with acute medical conditions.

**Methods:** Delirium assessment using the Confusion Assessment Method (CAM) upon admission and on day one, two and three or until delirium had resolved. Patients’ charts were reviewed if CAM was missing. In an linear mixed-effects model, the delirium incidence rate in AGCH was compared to pooled delirium incidence rates from six studies found in a high-quality review.

**Results:** 214 patients from the AGCH (mean age 81.9 years, 47% male, 12% with a history of dementia) were included in the analysis. Delirium developed in 8% (18/214) (95% confidence interval [CI] 5-13%) of patients during AGCH admission compared to 16% (95% CI 12-21%) in hospitals. Admission to the AGCH was associated with a decreased delirium incidence rate compared to the hospital control group (OR[odds ratio]= 0.49, 95% CI 0.24-0.98, p-value=0.044).

**Conclusions and implications:** The delirium incidence in the AGCH was low compared to those incidences found in general hospitals in literature. Based on these findings a controlled observational or randomized study measuring delirium in this care setting is recommended.

## Introduction

A common complication of hospitalization in older adults is the development of delirium, an acute disturbance in attention and cognitive functions.^1^ The etiology of delirium is considered multifactorial.^2^ Delirium is associated with negative health outcomes, including functional and cognitive decline, institutionalization, and mortality.^3,4^ The prevalence and incidence of delirium varies between settings and populations, with new-onset delirium during hospitalization ranging from 10% to 56%. ^5^

An alternative to conventional hospitalization is admission to an acute geriatric unit outside of a general hospital. This unit may be better adapted to the needs of older adults.^6^ In the Netherlands, the Acute Geriatric Community Hospital (AGCH) was introduced in 2018.^7^ This geriatrician-led unit located in a skilled nursing facility integrates specialized medical treatment with geriatric nursing care. This is the first unit of its kind in the Netherlands but other examples exist internationally.^8^ At the AGCH a non-pharmacological multi-component delirium prevention strategy has been implemented, consisting of encouraging early mobilization, preventing overstimulation (single rooms, noise reduction), management of delirium-inducing drugs and improving orientation through e.g. family involvement.^6,7^ It is unknown what the effect of this intervention is on the incidence of delirium in this new care setting.^7^ A feasibility study can help to determine if a large effectiveness study regarding delirium incidence at the AGCH should be conducted.^9^

We hypothesize that the non-pharmacological interventions at the AGCH reduce the incidence of delirium compared to usual care. The objective of this feasibility study was therefore to determine the incidence of delirium and compare this incidence to those incidences found in literature from general hospitals. This should help determine if an effect form this intervention in this new care setting is to be expected; and therefore determine if a larger prospectively controlled or randomized study on the incidence of delirium in the AGCH is advisable.

As secondary aims, we determined the duration of delirium and we quantified the use of pharmacological delirium treatment. The duration of delirium is relevant as it can also be shortened by a multi-component non-pharmacological intervention.^10^ Moreover, it is clinically relevant to know if patients (with or without delirium) were prescribed antipsychotics and/or benzodiazepines for the pharmacological treatment or prevention of delirium, as this is not recommended for the prevention of delirium.^11-13^

## Methods

### Design and setting

Data from a prospective cohort study were used. The study protocol was published elsewhere.^7^ Data collection started in February 2019 and was ceased in March 2020 during the COVID-19 pandemic.

Patients seen at the emergency department (ED) of the Amsterdam University Medical Centers in Amsterdam were assessed by an on-call geriatrician. Patients admitted to the AGCH were 65 years or older, presenting with an acute medical problem requiring hospitalization and one or more geriatric conditions, such as a fall, functional impairment or polypharmacy.^14^ Patients who did not require hospitalization, but needed short-term residential care in a skilled nursing facility, were excluded from admission to the AGCH. See the study protocol^7^ and appendix 1 for complete admission eligibility criteria.

### Ethical considerations

The local Ethics Committee of the the Amsterdam UMC, location AMC waived the obligation for the study to undergo formal ethical approval as described under Dutch law. We included patients who, or whose legal representative, could provide written informed consent. The study was registered in the Dutch Trial Registry, trial registration number NL7896.

### Control population from literature

We did not recruit a control group during the study period and we did not have delirium measurements available in a historical control group.^7^ To determine if a larger prospectively controlled study would be advisable we compared the incidence of delirium at the AGCH to existing literature. We searched for sources of aggregated data on the incidence rate of delirium in medical or geriatric (non-surgical) inpatients with a mean age of about 80 years (search strategy and excluded studies-appendix 2 and 3). We selected six studies from a review by Inouye et al. as a control group.^5^

### Measurement of incident delirium

Incident delirium, the number of new cases of delirium during admission, was the study outcome.^15^ No sample size was calculated. Patients were excluded from our analysis if delirium was present at the ED. The diagnosis of delirium was made by the geriatrician or geriatric nurse specialist by clinical assessment and using the Confusion Assessment Method (CAM).^16^ The CAM was filled out upon presentation to the ED and during the first three days of admission or until delirium had resolved. Nurses screened for signs of possible delirium, three times a day, during the first three days of admission using the Delirium Observation Screening Scale (DOSS).^17^ Patients were assessed by the same clinician for several consecutive days to recognize changes in mental status. On the weekend an on-call geriatrician assessed delirium status if delirium was clinically suspected. The DOSS and nursing chart covering the previous 24 hours were also considered in the delirium assessment. If there was a possible delirium after day three of admission, CAM assessments were continued until delirium had resolved.

### Duration of delirium

The duration of delirium was counted from the day the diagnosis was made until the CAM was permanently negative and/or the treating physician stated the delirium had resolved. In patients with an unresolved delirium at the time of discharge, we defined the first day of delirium until discharge as the duration of delirium.

### Use of antipsychotics and/or benzodiazepines

The administration of haloperidol, other antipsychotics, and benzodiazepines was collected from patients’ charts. We also checked if patients categorized as not delirious had received antipsychotics. This was 1) a check to see if no patient with a delirium diagnosis was missed and 2) a measure to quantify the use of antipsychotics and/or benzodiazepines as a preventive measure for delirium, although this is not recommended.^11-13^

### Statistical analysis

Descriptive statistics, chi-square, t-test, and Mann-Whitney U test were used to compare patients with and without delirium upon admission. To compare incidence rates from literature we pooled studies in a meta-analysis of proportions, using a random-effects model.^18^ We tested if the difference in delirium incidence was statistically significant by creating a logistic mixed-effects meta-regression model with the location of the study (hospital versus AGCH) as a moderator.^19^ We did not perform meta-regression of other covariates because the number of included studies was limited (<10).^18^ All analyses were performed using SPPS version 26.00 (IBM SPSS Statistics, IBM Corporation, Armonk, NY) and R version 3.6.1. We used the metaphor (Viechtbauer, 2010) and meta (Schwarzer et al., 2015) packages in R.

## Results

Between January 31, 2019 and March 13, 2020, a total of 466 consecutive patients were admitted to the AGCH (figure 1). Of the 261 patients who participated in the study 47 were excluded because of prevalent delirium or because of missing delirium assessments at the ED. The sample for this study therefore consisted of 214 patients (figure 1). Mean (SD) age was 81.9 (8.1) years, 47.2% was male, 12.1% had a diagnosis of dementia, and 47.2% of the patients was frail (table 1). Development of delirium during admission occurred in 18 out of 214 patients, which is an incidence rate of 8.4% (95% CI [confidence interval] 5-13%). The median (IQR [interquartile range]) duration of delirium in the AGCH was 2.5 days (1.0-5.3) (table 1). Mean length of stay (SD) was 9.6 (7.3) days in all patients, 9.4 (7.4) days in patients with no delirium and 11.9 (6.4) days in patients with delirium. Median length of stay (IQR) was 7.0 (5.0-11.00) days in patients with no delirium and 10 (7.5-16.8) days in patients with delirium.

**Table 1.**
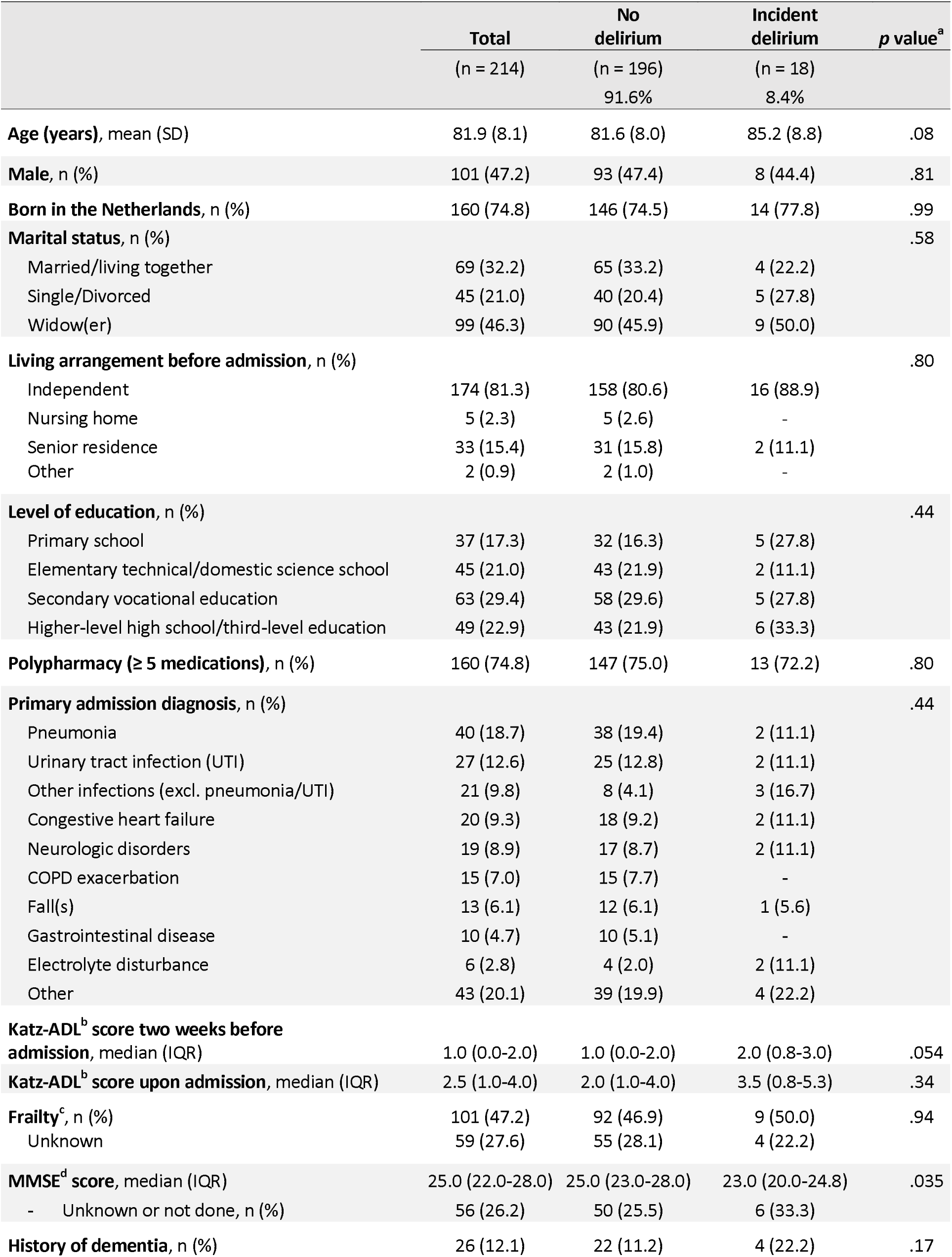
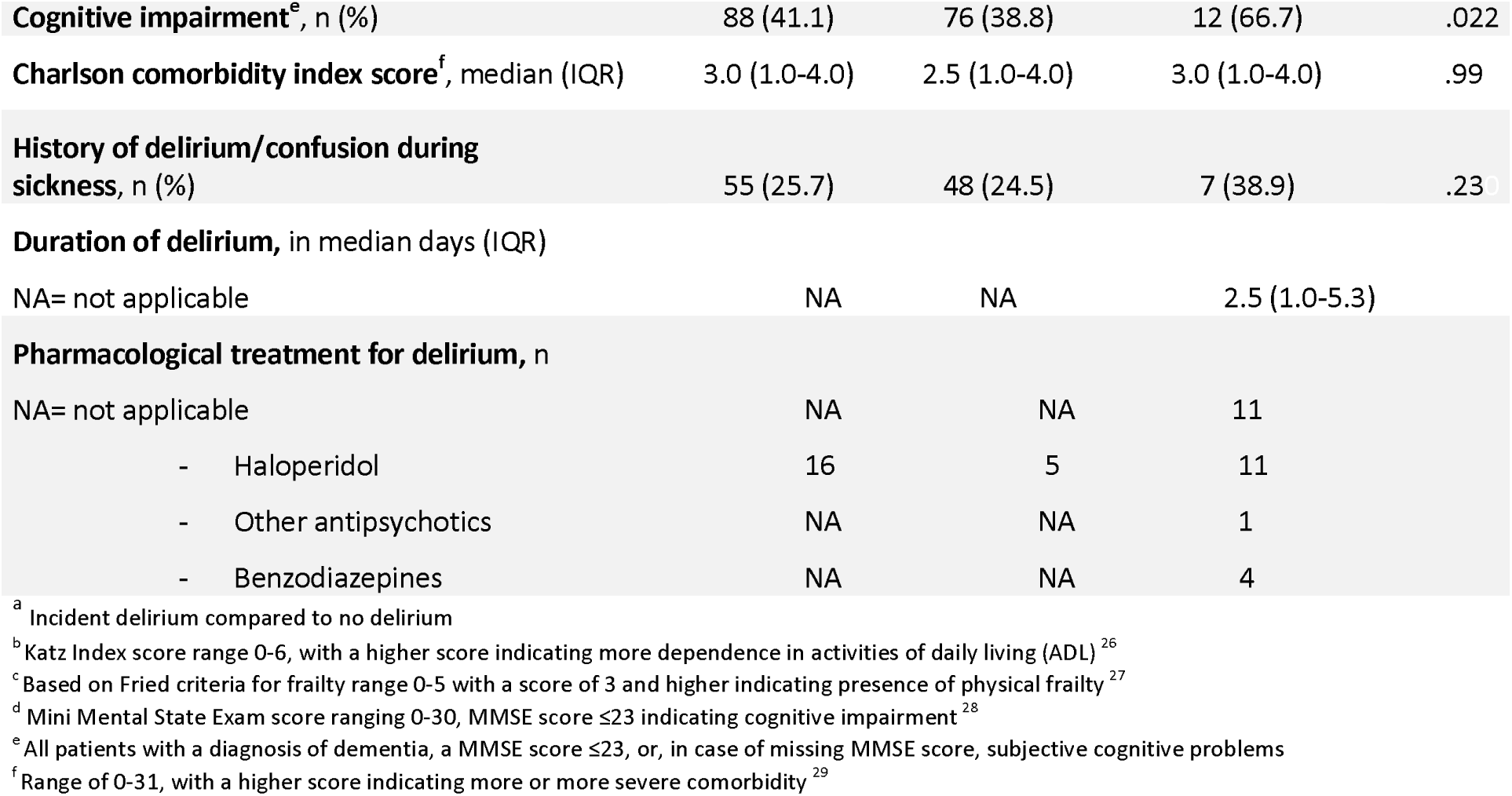
Baseline characteristics of the total study population grouped by patients with and without delirium.

**Figure 1.**
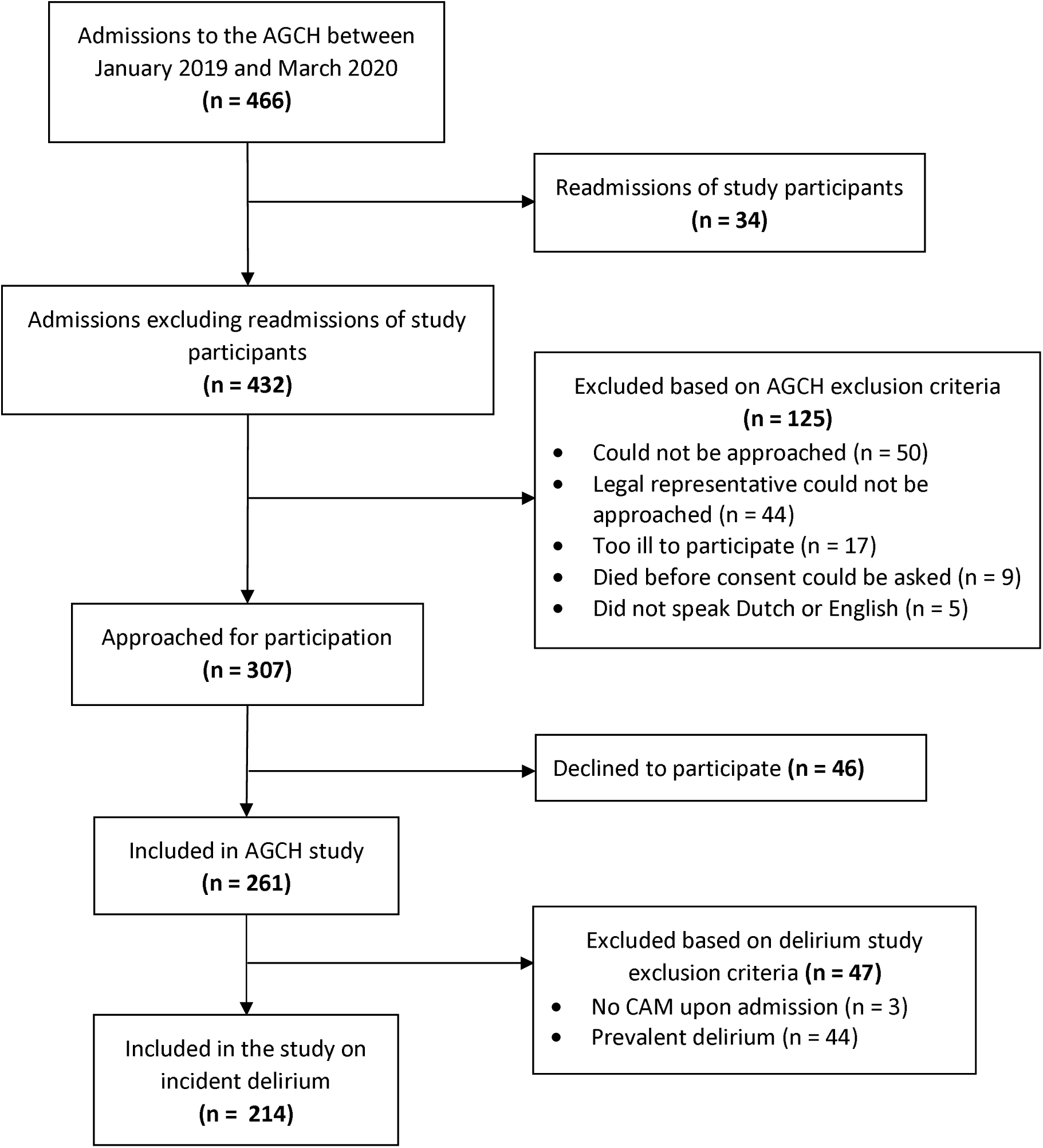
Participant flow-chart Acute Geriatric Community Hospital (AGCH) study.

### Pharmacological treatment for delirium

Eleven out of 18 patients (61.1%) with a diagnosis of delirium were administered medication for the treatment of delirium. Haloperidol was administered most frequently (n=11). The regular prescription of haloperidol was 0.5-2.0mg per dose, typically given once a day, or twice in case of severe delirium, with a maximum of three dosages. Five (5 out of 196, 2.6%) patients without delirium were administered haloperidol, either as prevention due to a high risk of delirium or as treatment for pre-existing symptoms unrelated to delirium (table 1).

### Delirium incidence in comparison to reference group from literature

The control group was based on six studies (appendix 4).^5^ In total 1546 study participants with a mean age of 80 years (Appendix 4). None of the studies, except for Friedman et al.^20^ reported to have implemented multi-component delirium prevention strategies, we therefore assumed usual care was delivered. The pooled delirium incidence rate of these six studies was 16% (95% CI random effects model 12-21%) (Figure 2). The meta-analysis showed a high heterogeneity (I^2^=84%). In a separate logistic mixed-effects model comparing general hospitals (reference category) versus the AGCH, we found that admission to the AGCH was associated with a decrease in delirium incidence (OR [odds ratio]= 0.49, 95% CI 0.24-0.98, p=.044).

**Figure 2.**
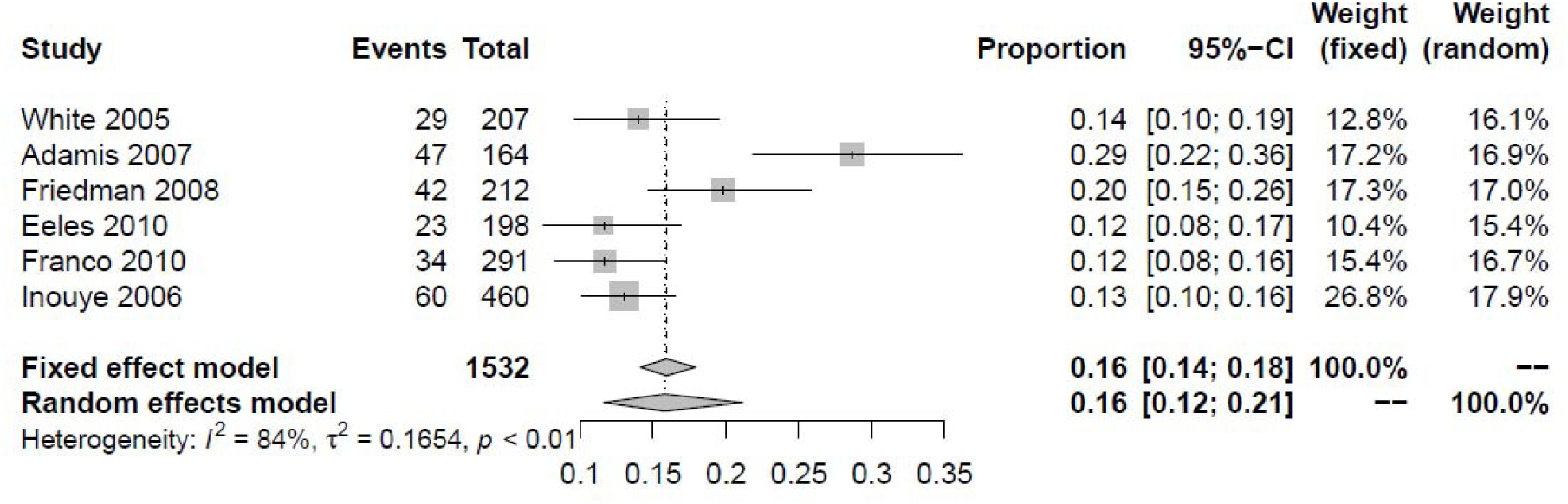
Meta-analysis of proportions of delirium incidences in older hospitalized medical patients found in literature^5^. The pooled incidence rate of these six studies was 16% (95% CI [confidence interval]random effects model 12-21%).

### Adherence to CAM evaluations and missing data

In patients with delirium 27.8% of total CAM evaluations and 46.3% of total DOSS scores were missing during the first three days of admission. For patients without delirium 46.9% and 66.7% were missing, respectively. In 15% of all cases all three CAM evaluations were missing. Based on the CAM evaluation and daily delirium assessment by the attending clinician we could ascertain the presence delirium in the first three days of admission in all patients.

## Discussion

We measured the effect of a non-pharmacological multi-component delirium prevention strategy at the AGCH and found an incidence rate of delirium of 8.4%. This incidence is lower compared to rates found in hospital medical or geriatric wards found in historical cohorts from literature. This finding is in line with previous literature on multi-component interventions for preventing delirium in hospitalized patients: a 2016 Cochrane review reports moderate quality evidence that multi-component interventions in medical, non-surgical, patients lower delirium incidence.^6^ Moreover, the median duration of delirium of 2.5 days at the AGCH is comparable to the duration that is found in literature on non-pharmacological interventions.^21^ The prescription rate of medication (61.1%) may be lower in the AGCH compared to other studies, which report rates of 74-86%.^22,23^ The Dutch guideline on delirium, and international guidelines alike, recommends to take a cautious approach to the prescription of medication for the treatment of delirium.^11-13^ In addition, only a few patients received medication, in this case haloperidol, for the prevention of delirium, meaning that there were not many non-delirious patients receiving haloperidol. This is relevant because, administration of anti-psychotics such as haloperidol could lower delirium incidence rates in high incidence groups.^11^ Moreover, not all CAM measurement on day 1-3 of admission were complete, but it was possible to ascertain the presence of delirium based on daily clinical delirium assessment.

A strength of this feasibility study is the relatively large study sample. Limitations of the study include that the incidence rate of delirium could have been influenced by selection bias as legal representatives of patients could not always be contacted to obtain consent. Moreover, even though we selected a control group from a high-quality review article; this review was not recently published (2014) and the selected studies were conducted in different countries than the Netherlands.^5^ We also did not have insight into all of the baseline characteristics of these studies, which makes it difficult to assess comparability. In addition, we could not definitively ascertain that ‘usual care’ was delivered in each unit or what this was composed of. Finally, we did not collect data on illness severity, which can be associated with delirium.^24^

## Conclusion and implications

This feasibility study shows that the incidence rate of delirium in the AGCH may be lower than in general hospitals. Based on this result we would recommend a randomized controlled study or a two-armed observational study using e.g. inversely weighted propensity scores ^25^ to test if admission to the AGCH is effective in reducing the incidence of delirium. Moreover, attention should be given to collecting complete CAM assessments in this ‘real-world’-setting. If in a larger, prospective and controlled study the incidence of delirium at the AGCH is lower than in hospital this would support the implementation of the AGCH model of care elsewhere.

## Data Availability

Data is currently not available for reuse

## Acknowledgements

This work was supported by ZonMw, the Netherlands Organization for Health Research and Development [grant number 808393598041] and the PVE- fund.. Moreover, the care provided at the AGCH is provided in a partnership between Cordaan, a community and home-care organization and the Amsterdam University Medical Center, location Academic Medical Center The AGCH is financially supported by Zilveren Kruis, a health insurance company.

The authors thank all professionals who have worked on the development of the AGCH. Also, we would like to thank the members of the AGCH study group, these are the clinicians who work at the Geriatrics Department of the Amsterdam University Medical Centers and who support the data collection at the AGCH. We would also like to thank Marije Wolvers for her advice on the statistical analysis.

## References

1. American Psychiatric Association. Diagnostic and Statistical Manual of Mental Disorders, Fifth Edition. Arlington, VA2013.

2. Inouye SK, Charpentier PA. Precipitating factors for delirium in hospitalized elderly persons. Predictive model and interrelationship with baseline vulnerability. Jama. 1996;275(11):852–857.

3. Inouye SK, Rushing JT, Foreman MD, Palmer RM, Pompei P. Does delirium contribute to poor hospital outcomes? A three-site epidemiologic study. Journal of general internal medicine. 1998;13(4):234–242.

4. Fick DM, Agostini JV, Inouye SK. Delirium superimposed on dementia: a systematic review. Journal of the American Geriatrics Society. 2002;50(10):1723–1732.

5. Inouye SK, Westendorp RG, Saczynski JS. Delirium in elderly people. Lancet (London, England). 2014;383(9920):911–922.

6. Siddiqi N, Harrison JK, Clegg A, et al. Interventions for preventing delirium in hospitalised non-ICU patients. The Cochrane database of systematic reviews. 2016;3:Cd005563.

7. Ribbink ME, Macneil-Vroomen JL, van Seben R, Oudejans I, Buurman BM. Investigating the effectiveness of care delivery at an acute geriatric community hospital for older adults in the Netherlands: a protocol for a prospective controlled observational study. BMJ open. 2020;10(3):e033802.

8. Ribbink ME, Gual N, MacNeil-Vroomen JL, et al. Two European Examples of Acute Geriatric Units Located Outside of a General Hospital for Older Adults With Exacerbated Chronic Conditions. Journal of the American Medical Directors Association. 2021;22(6):1228–1234.

9. Bowen DJ, Kreuter M, Spring B, et al. How we design feasibility studies. Am J Prev Med. 2009;36(5):452–457.

10. Inouye SK, Bogardus ST, Jr., Charpentier PA, et al. A multicomponent intervention to prevent delirium in hospitalized older patients. The New England journal of medicine. 1999;340(9):669–676.

11. Nederlandse Vereniging voor Klinische Geriatrie. Richtlijn Delier Volwassenen. 2013.

12. Excellence NIfHaC. Delirium: prevention, diagnosis and management. https://www.nice.org.uk/guidance/cg103/chapter/1-Guidance#interventions-to-prevent-delirium. Published 2010. Updated March 2019. Accessed 10th of March 2021.

13. American Geriatrics Society Expert Panel on Postoperative Delirium in Older A. American Geriatrics Society abstracted clinical practice guideline for postoperative delirium in older adults. Journal of the American Geriatrics Society. 2015;63(1):142–150.

14. Buurman BM, Hoogerduijn JG, de Haan RJ, et al. Geriatric conditions in acutely hospitalized older patients: prevalence and one-year survival and functional decline. PloS one. 2011;6(11):e26951.

15. National Institute for Health and Care Excellence. Delirium: Diagnosis, Prevention and Management. London 2010.

16. Inouye SK, van Dyck CH, Alessi CA, Balkin S, Siegal AP, Horwitz RI. Clarifying confusion: the confusion assessment method. A new method for detection of delirium. Annals of internal medicine. 1990;113(12):941–948.

17. Schuurmans MJ, Shortridge-Baggett LM, Duursma SA. The Delirium Observation Screening Scale: a screening instrument for delirium. Research and theory for nursing practice. 2003;17(1):31–50.

18. Wang N. How to Conduct a Meta-Analysis of Proportions in R: A Comprehensive Tutorial. 2018.

19. Viechtbauer W. Comparing Estimates of Independent Meta-Analyses or Subgroups. https://www.metafor-project.org/doku.php/tips:comp_two_independent_estimates. Published 2020. Accessed.

20. Friedman SM, Mendelson DA, Bingham KW, McCann RM. Hazards of hospitalization: residence prior to admission predicts outcomes. The Gerontologist. 2008;48(4):537–541.

21. Martinez FT, Tobar C, Beddings CI, Vallejo G, Fuentes P. Preventing delirium in an acute hospital using a non-pharmacological intervention. Age and ageing. 2012;41(5):629–634.

22. van Velthuijsen EL, Zwakhalen SMG, Mulder WJ, Verhey FRJ, Kempen G. Detection and management of hyperactive and hypoactive delirium in older patients during hospitalization: a retrospective cohort study evaluating daily practice. International journal of geriatric psychiatry. 2018;33(11):1521–1529.

23. Nguyen PV-Q, Malachane A, Minh TTV. Antipsychotic prescription patterns in the management of delirium symptoms in hospitalized elderly patients. Proceedings of Singapore Healthcare. 2017;26(4):230–234.

24. Inouye SK, Viscoli CM, Horwitz RI, Hurst LD, Tinetti ME. A predictive model for delirium in hospitalized elderly medical patients based on admission characteristics. Annals of internal medicine. 1993;119(6):474–481.

25. Austin PC, Stuart EA. Moving towards best practice when using inverse probability of treatment weighting (IPTW) using the propensity score to estimate causal treatment effects in observational studies. Stat Med. 2015;34(28):3661–3679.

26. Katz S, Ford AB, Moskowitz RW, Jackson BA, Jaffe MW. Studies of illness in the aged. The index of ADL: A standardized measure of biological and pshychosocial function. Jama. 1963;185:914–919.

27. Fried LP, Ferrucci L, Darer J, Williamson JD, Anderson G. Untangling the concepts of disability, frailty, and comorbidity: implications for improved targeting and care. The journals of gerontology Series A, Biological sciences and medical sciences. 2004;59(3):255–263.

28. Folstein MF, Folstein SE, McHugh PR. “Mini-mental state”. A practical method for grading the cognitive state of patients for the clinician. Journal of psychiatric research. 1975;12(3):189–198.

29. Charlson ME, Pompei P, Ales KL, MacKenzie CR. A new method of classifying prognostic comorbidity in longitudinal studies: development and validation. Journal of chronic diseases. 1987;40(5):373–383.

