## Appendix 1-4 for "The incidence of delirium in an acute geriatric community hospital: an observational cohort feasibility study"

**Appendix 1 –** Admission criteria Acute Geriatric Community Hospital (AGCH).

*Criteria upon assessment at the emergency department:*
1) Acute medical problems in older patients that require hospitalization, e.g., acute events such as a pneumonia or an exacerbation of chronic conditions such as heart failure or minor acute events in very frail patients.

2) Hemodynamic stability.

3) No need for complex diagnostic testing such as CT- or MRI scans during admission.

4) Expecting to return to previous living situation in 14 days.

5) Geriatric conditions e.g., delirium, cognitive impairment, falls and/or functional impairment.

**Appendix 2** – Search strategy studies reporting incident delirium in medical inpatients.

Methods

We looked for sources of aggregated data (guidelines) or reviews reporting the incidence of delirium in older adults admitted to acute medical units and extracted the data from the individual studies that reported in the guideline.

Secondly, we performed a search on PubMed looking for (systematic) reviews or meta-analyses reporting incidence of delirium in older adults admitted to acute medical units.

*Search strategy in PubMed*

((delirium[MeSH Terms]) AND ((internal medicine[MeSH Terms])OR(medical[Title/Abstract])) NOT (surgical[Title/Abstract]) NOT (orthopeadic[Title/Abstract]) NOT (hip fracture[Title]) NOT (fracture[Title]) NOT (pediatric[Title/Abstract]) NOT (postoperative[Title/Abstract]))

Filters applied: Meta-Analysis, Systematic Review

Criteria for including studies were:

*Inclusion criteria*

- *Older adults aged 60 years and older*
- *Admitted to medial or geriatric wards of acute or general hospitals*
- *Prospective cohort and cross-sectional studies*
- *Control arms of clinical trials*

*Exclusion criteria*

- *Mean age study population <65 years*
- *Including data from intervention arm of clinical trials*
- *Case-control studies*
- *Retrospective studies*
- *Data from surgical wards or intensive care units*

**Appendix 3** – found and excluded studies.

*Clinical guideline on delirium and or review:*

1) **Excluded:** **NICE guideline delirium 2010 ^1^** We found a guideline reporting the incidence of delirium in general medicine wards; the NICE guideline for delirium which reports an incidence of 15.2% (range 12.5-17.9) based on two studies.^1^ One of these studies included the intervention arm of a clinical trial^2^ and therefore we decided not to use the NICE guideline report on incident delirium as a control group.

2) **Included:** **Inouye et al. 2014 ^3^-** This is the high-quality review from which we used included studies as a control group.

*Results from PubMed search looking for systematic reviews or meta-analyses:*

Our search yielded 21 hits (1,124 without filters) in which we found two relevant systematic reviews based on screening title and abstract:

1) **Excluded:** **Siddiqi et al. 2006 ^4^** We found a systematic review by Siddiqi et al.^4^ that reported on the incidence of delirium in 14 studies, due to the smaller sample sizes and studies being published before 2005 we decided not to use this systematic review.

2) **Excluded**: **Ahmed et al. 2014 ^5^** We found a systematic review by Ahmed et al.,^5^ including 11 studies that report delirium incidence in older medical patients, but in this study there was a risk of bias because it aimed to describe risk factors for delirium and not solely the incidence of delirium. This meant case-control studies were used and possible relevant studies were excluded. Therefore, we decided not to use this review.

**Flow-chart of selection of studies (aggregated data and (systematic reviews) for hospital control group :**

Sources of aggregated data and/or reviews (n=2)

+ PubMed search (n=21)

**n=23**

Excluded from search based on title and abstract (n=19)

*Aggregated data/reviews*:

NICE guideline 2010

Inouye et al. 2014

*From PubMed search:*

Siddiqi et al. 2006

Ahmed et al. 2014

Ahmed et al. 20014

Ahme

Excluded (n=3)

- Including intervention study (NICE guideline 2010)
- Low sample sizes and less recent studies compared to Inouye et al. 2014 (Siddiqi et al.2006)
- Including case-control studies and potential bias because of study aim (Ahmed et al. 2014)

Inouye et al. 2014, see table 2 for included studies

**Appendix 4 table** –Studies included in meta-analysis of proportions of delirium incidence, studies from a 2014 review on delirium incidence by Inouye et al. CAM, Confusion Assessment Method; DRS, Delirium Rating Scale; DSM–IV, Diagnostic and Statistical Manual of Mental Disorders (4rd Ed). * original sample included prevalent delirium

| Study (authors, publication year, country) | Study setting | Study design | Patients with delirium (n) | Patients without delirium  (n) | Total sample (n) | Incidence rate (%) | Age, years | Mean age | Sex: male/female | Criteria for delirium | Assessment frequency (h) |
| --- | --- | --- | --- | --- | --- | --- | --- | --- | --- | --- | --- |
| Franco *et al.,* 2010, Colombia ^6^ | Medical | Case control | 34 | 257 | 291 | 12% | ≥59 | 74 | 105/186 | CAM,DRS | 24 |
| White 2005, United Kingdom ^7^ | Medical | Cohort | 29 | 178 | 207* | 14% | ≥75 | 82 | Only number of males in total group | DAM,DSMIV | 24 |
| Adamis 2007, United Kingdom ^8^ | Geriatric | Cohort | 47 | 117 | 164 | 29% | ≥70 | 85 | 54/110 | CAM,DRS | 72 |
| Friedman 2008, United States ^9^ | Geriatric | Cohort | 42 | 171 | 212 | 20% | ≥65 | 79 | 85/127 | CAM | 24 |
| Eeles 2010, United Kingdom ^10^ | Medical | Cohort | 23 | 175 | 198* | 11% | ≥74 | 83 | 117 males in total group | DSMIV | 48 |
| Inouye et al. 2006, United States ^11^ | Medical | Cohort | 60 | 400 | 460 | 13% | ≥70 | 80 | 183/277 | CAM | 24 |

**References to appendix**

1.	National Institute for Health and Care Excellence. *Delirium: Diagnosis, Prevention and Management.* London2010.

2.	Leslie DL, Marcantonio ER, Zhang Y, Leo-Summers L, Inouye SK. One-year health care costs associated with delirium in the elderly population. *Archives of internal medicine.* 2008;168(1):27-32.

3.	Inouye SK, Westendorp RG, Saczynski JS. Delirium in elderly people. *Lancet (London, England).* 2014;383(9920):911-922.

4.	Siddiqi N, House AO, Holmes JD. Occurrence and outcome of delirium in medical in-patients: a systematic literature review. *Age and ageing.* 2006;35(4):350-364.

5.	Ahmed S, Leurent B, Sampson EL. Risk factors for incident delirium among older people in acute hospital medical units: a systematic review and meta-analysis. *Age and ageing.* 2014;43(3):326-333.

6.	Franco JG, Valencia C, Bernal C, et al. Relationship between cognitive status at admission and incident delirium in older medical inpatients. *J Neuropsychiatry Clin Neurosci.* 2010;22(3):329-337.

7.	White S, Calver BL, Newsway V, et al. Enzymes of drug metabolism during delirium. *Age and ageing.* 2005;34(6):603-608.

8.	Adamis D, Treloar A, Darwiche FZ, Gregson N, Macdonald AJ, Martin FC. Associations of delirium with in-hospital and in 6-months mortality in elderly medical inpatients. *Age and ageing.* 2007;36(6):644-649.

9.	Friedman SM, Mendelson DA, Bingham KW, McCann RM. Hazards of hospitalization: residence prior to admission predicts outcomes. *The Gerontologist.* 2008;48(4):537-541.

10.	Eeles EM, Hubbard RE, White SV, O'Mahony MS, Savva GM, Bayer AJ. Hospital use, institutionalisation and mortality associated with delirium. *Age and ageing.* 2010;39(4):470-475.

11.	Inouye SK, Zhang Y, Han L, Leo-Summers L, Jones R, Marcantonio E. Recoverable cognitive dysfunction at hospital admission in older persons during acute illness. *Journal of general internal medicine.* 2006;21(12):1276-1281.
